# A case-control study based on the National Health and Nutrition Examination Survey to evaluate the effects of human papilloma virus on bone health in women

**DOI:** 10.1101/2022.09.13.22279899

**Authors:** Xiang Li, Guangjun Jiao, Yunzhen Chen

**Affiliations:** Shandong University Cheeloo College of Medicine, Jinan, Shandong, China, 250000; Qilu Hospital of Shandong University, Department of Spine Surgery, Jinan, Shandong, China, 250000

**Keywords:** Women, Human papillomavirus, Bone mineral density, Correlation

## Abstract

**Background:** Globally, both human papillomavirus (HPV) infection and osteoporosis (OP) are more prevalent in women than in men. It remains unclear whether HPV has an impact on bone health.

**Methods:** This case-control study was based on data from the National Health and Nutrition Examination Survey (NHANES). Comparable datasets were created via the nearest neighbor propensity score matching (PSM) method (1:2). The Welch two-sample t test was used to analyze the association between HPV infection and bone mineral density (BMD). Restricted cubic spline (RCS) and Kendall’s tau-b tests were used to explore the effect of HPV infection type on BMD.

**Results:** BMDs in the legs and lumbar spine were lower in subjects infected with HPV than in uninfected subjects. RCS analysis showed that the larger the number of cooccurring HPV types in a woman, the lower the BMD was. In addition, four HPV types were negatively associated with leg BMD, and 14 HPV types were negatively associated with lumbar spine BMD. HPV types 53, 59, and 89 had effects on both leg and lumbar spine BMDs.

**Conclusions:** HPV infection was associated with a decrease in BMD. Appropriately designed trials can help determine whether interventions to prevent HPV infection will have a protective effect on BMD.

**Funding:** This research was not supported by any specific grant from any funding agency in the public, commercial or not-for-profit sector.

## 1. Introduction

Osteoporosis (OP) is one of the most common chronic diseases of the skeleton. It is characterized by a decrease in bone mineral density (BMD) and the destruction of bone structure[1, 2]. OP is of great clinical importance because OP is a major risk factor for fracture and greatly affects human health[3, 4]. It has been reported that there are more than 200 million OP patients worldwide[5, 6]. In some regions, the prevalence of OP is approximately 40% in the elderly population[7-9]. OP and associated fractures impose substantial economic and social burdens. OP, a complex disease, is associated with a variety of factors. A better understanding of emerging risk factors contributing to the disease may enable the development of prevention and treatment strategies targeting high-risk individuals.

Human papillomavirus (HPV) is one of the most recognized viruses in humans, with a fairly high infection rate (2-44%)[10-12]. Most individuals are exposed to HPV infection at some point in their lifetime. HPV resulting in lesions on the genitalia has become the most common sexually transmitted infection in the world. Moreover, HPV is also recognized as a causative factor for cervical cancer[13]. It is also a risk factor for several tumors[14-19]. The rate of HPV infection is significantly higher among women than among men[20, 21]. This is similar to OP, for which female sex is a risk factor[22-24]. However, the role of HPV in the bone health of women is unclear, and no studies have confirmed the association between HPV and BMD.

Here, we conducted a large case-control study based on National Health and Nutrition Examination Survey (NHANES) to explore the association between HPV and BMD. Elucidating the strength and pattern of this association will provide evidence to guide appropriate targeted public health interventions.

## 2. Subjects and methods

### 2.1 General information about the NHANES

The National Center for Health Statistics of the Centers for Disease Control and Prevention in the United States conducted a multistage, stratified, large-scale, nationally representative study named the NHANES[25]. This cross-sectional survey collected demographic, dietary, examination, and questionnaire data. The NHANES protocol is described in detail elsewhere[26]. In this study, the data were obtained from the 2009–2014 NHANES database.

### 2.2 Definition of variables

#### 2.2.1 HPV infection

All eligible female participants underwent vaginal swab sample collected. The obtained samples were tested with the Roche linear array HPV genotyping test to determine the presence of HPV infection and the type of HPV in cases of infection. This method can detect 37 HPV types: 6, 11, 16, 18, 26, 31, 33, 35, 39, 40, 42, 45, 51, 52, 53, 54, 55, 56, 58, 59, 61, 62, 64, 66, 67, 68, 69, 70, 71, 72, 73, 81, 82, 83, 84, 89, and IS39.

We included subjects who were not infected with HPV in group A and those who were infected (all types) into group B. Subjects with missing data were excluded.

#### 2.2.2 Measurement of BMD

Dual energy X-ray absorptiometry was performed to detect the BMD of eligible female subjects. The scanning area included the whole body. However, in this study, we focused on only the BMD of the subjects’ legs and lumbar spines.

For the data from 2009 to 2010, the BMD of the leg was the average of bilateral total femur BMD, and the BMD of the lumbar spine was the average of L1–4 BMD. For the data from 2011 to 2014, the BMD value of the leg was the average BMD in both legs. In addition, if the BMD value of one leg was missing, the BMD in that leg was considered the value of the other leg. Subjects for whom the BMD in the legs or lumbar spine could not be calculated were excluded.

#### 2.2.3 Covariates

We included the following covariates of interest: age, race, education level, marital status, poverty-to-income ratio, smoking status, alcohol consumption status, diabetes, depression score and sleeping difficulty.

Smoking was defined as smoking ≥ 100 cigarettes in a lifetime[27]. Alcohol consumption was defined as having 4/5 or more drinks every day[28, 29].

Subjects were considered to have diabetes if they met one of the following criteria: told by doctors or other health professionals they had diabetes, insulin use, or diabetes medication use to lower their blood sugar[30].

Subjects with a Patient Health Questionnaire (PHQ)-9 total score of ≥ 10 were considered to have clinically relevant depression[8, 31]. Subjects who answered “yes” to the question “Have you ever been told by doctor that you have a sleep disorder?” were considered to have sleeping difficulty.

### 2.3 Ethical statement

The National Center for Health Statistics Research Ethics Review Board had approved the NHAHES. Therefore, NHANES data could be accessed without ethical or administrative permission.

### 2.4 Statistical analyses

We used normal Q-Q plots to test the distribution of the data. Welch two-sample t tests or nonparametric Mann–Whitney U tests were used to analyze continuous variables. Chi-square tests or Fisher’s exact tests were used to compare categorical variables.

Propensity score matching (PSM) has been shown to be effective in reducing selection bias in retrospective studies[7, 32, 33]. We first transformed BMD into a categorical variable based on whether BMD was ≥1. Subsequently, PSM, using the 1:2 nearest neighbor matching algorithm, was used to match subjects.

In the case of nonlinear a priori associations, restricted cubic spline (RCS) models were used to describe dose-response relationships between the continuous exposure and outcomes[34, 35]. In this study, RCS with five knots was used to further explore the association between HPV infection and BMD.

Finally, Kendall’s tau-b test was used to analyze the association between HPV type and BMD. In this analysis, we calculated only the individual effects of each HPV type and did not consider interactions between different types. Since there were no subjects infected with only HPV type 26, 64 or 82, we could not analyze their effects. P < 0.05 was considered statistically significant, and all tests were one-sided. Data extraction and merging, statistical analyses and figures were completed with R (version 4.1.1).

## 3. Results

### 3.1 Participant characteristics before matching

After screening, a total of 4673 female subjects were included in the study. The mean age of group A (n = 2614) was 39 ± 11.97 years, and the mean age of group B (n = 2059) was 36.67 ± 12.29 years. Supplemental table 1 shows the comparisons of the baseline characteristics between the two groups. The results indicated statistically significant differences for the vast majority of characteristics (P < 0.001). This suggested significant selection bias between the two groups of subjects. Therefore, performed PSM to eliminate this phenomenon.

**Table 1.**
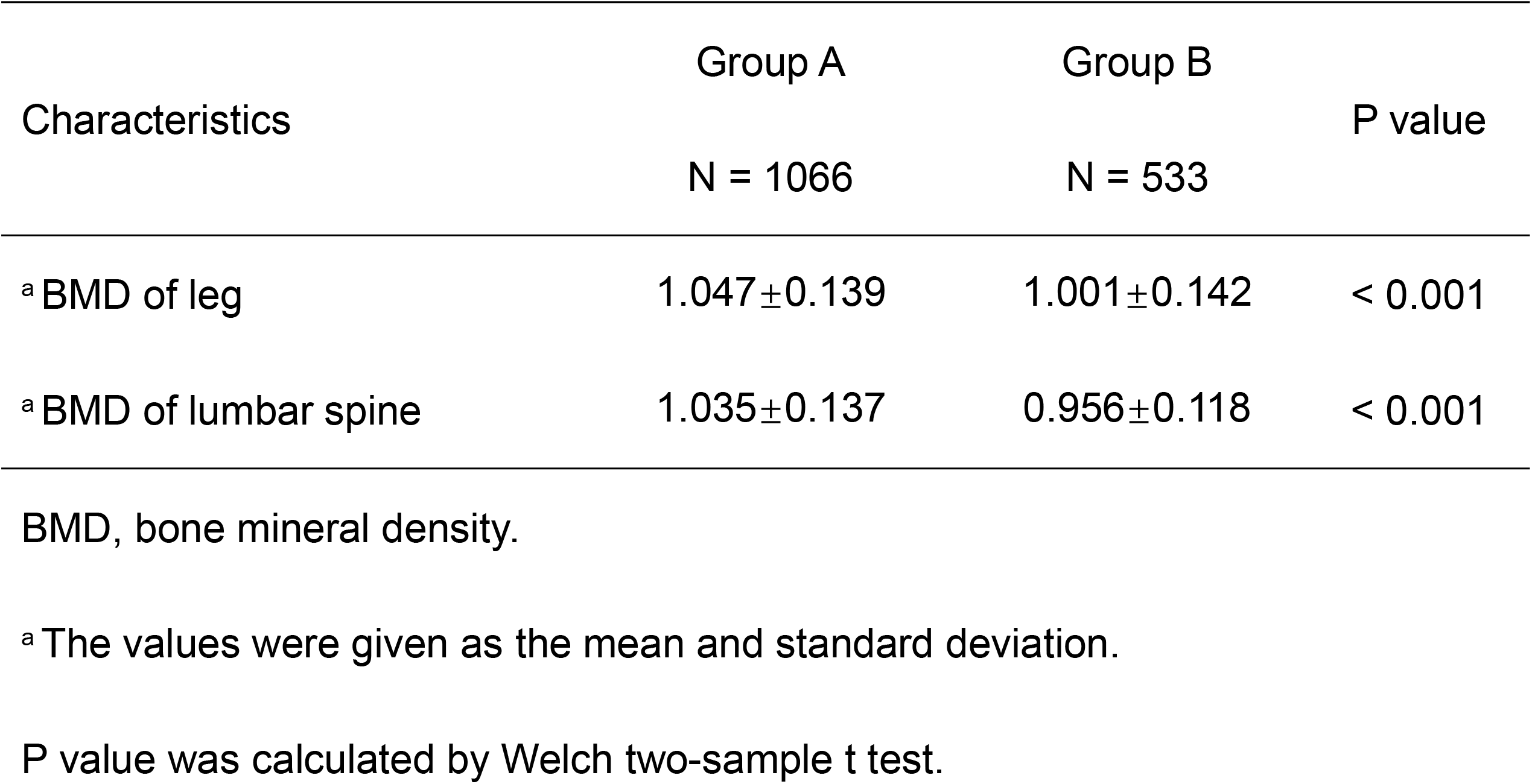
Comparison of BMD between the two groups after matching. BMD, bone mineral density.

### 3.2 Construction of matched datasets

After 1:2 matching, we obtained datasets for BMDs in the legs and lumbar spine. In total, 1066 subjects in group A and 533 subjects in group B were included. Analysis of baseline characteristics showed that the selection bias had been eliminated (P > 0.05). More detailed data are presented in Supplemental table 2 and Supplemental table 3. The BMD values of the legs and lumbar spine in the different age groups are shown in Fig. 1A. Normal Q-Q plots showed a significant normal distribution of BMD (Fig. 1B - D).

**Table 2.** The correlation between HPV types and BMD of leg.

| Types | Coefficient | P value | Types | Coefficient | P value | Types | Coefficient | P value |
| --- | --- | --- | --- | --- | --- | --- | --- | --- |
| 06 | -0.042 | .047 | 52 | -0.016 | .264 | 69 | 0.027 | .144 |
| 11 | - | - | 53 | -0.042 | .046 | 70 | 0.006 | .399 |
| 16 | -0.025 | .162 | 54 | 0.002 | .463 | 71 | 0.002 | .473 |
| 18 | -0.020 | .218 | 55 | -0.014 | .291 | 72 | -0.023 | .181 |
| 31 | -0.031 | .105 | 56 | 0.006 | .406 | 73 | 0.000 | .499 |
| 33 | -0.160 | .267 | 58 | 0.029 | .122 | 81 | -0.056 | .012 |
| 35 | 0.023 | .181 | 59 | -0.052 | .019 | 83 | 0.004 | .429 |
| 39 | -0.008 | .376 | 61 | 0.029 | .120 | 84 | -0.008 | .375 |
| 40 | -0.017 | .249 | 62 | -0.016 | .262 | 89 | -0.060 | .008 |
| 42 | 0.019 | .228 | 66 | -0.021 | .206 | IS39 | -0.009 | .353 |
| 45 | 0.025 | .159 | 67 | -0.001 | .488 |  |  |  |
| 51 | -0.011 | .331 | 68 | -0.013 | .298 |  |  |  |

BMD, bone mineral density.
P values were calculated by Kendall's tau-b correlation test.
HPV types that were statistically significant were marked in red.
Since there were no subjects infected with only HPV types 11, it was not possible to calculate their effects on the BMD of leg.

**Table 3.** The correlation between HPV types and BMD of lumbar spine.

| Types | Coefficient | P value | Types | Coefficient | P value | Types | Coefficient | P value |
| --- | --- | --- | --- | --- | --- | --- | --- | --- |
| 06 | -0.028 | .134 | 52 | -0.010 | .340 | 69 | -0.015 | .270 |
| 11 | -0.038 | .066 | 53 | -0.085 | < .001 | 70 | -0.068 | .003 |
| 16 | -0.050 | .022 | 54 | -0.029 | .120 | 71 | 0.013 | .309 |
| 18 | -0.033 | .094 | 55 | -0.055 | .014 | 72 | -0.051 | .021 |
| 31 | -0.034 | .086 | 56 | -0.040 | .055 | 73 | -0.044 | .038 |
| 33 | -0.075 | .001 | 58 | -0.040 | .056 | 81 | -0.065 | .004 |
| 35 | 0.000 | .496 | 59 | -0.049 | .024 | 83 | -0.048 | .028 |
| 39 | -0.011 | .328 | 61 | -0.017 | .246 | 84 | -0.056 | .012 |
| 40 | -0.024 | .167 | 62 | -0.057 | .010 | 89 | -0.046 | .033 |
| 42 | -0.029 | .120 | 66 | -0.029 | .122 | IS39 | 0.033 | .091 |
| 45 | -0.022 | .194 | 67 | -0.030 | .115 |  |  |  |
| 51 | -0.026 | .152 | 68 | -0.042 | .047 |  |  |  |

BMD, bone mineral density.
P values were calculated by Kendall's tau-b correlation test.
HPV types that were statistically significant were marked in red.

**Figure 1.**
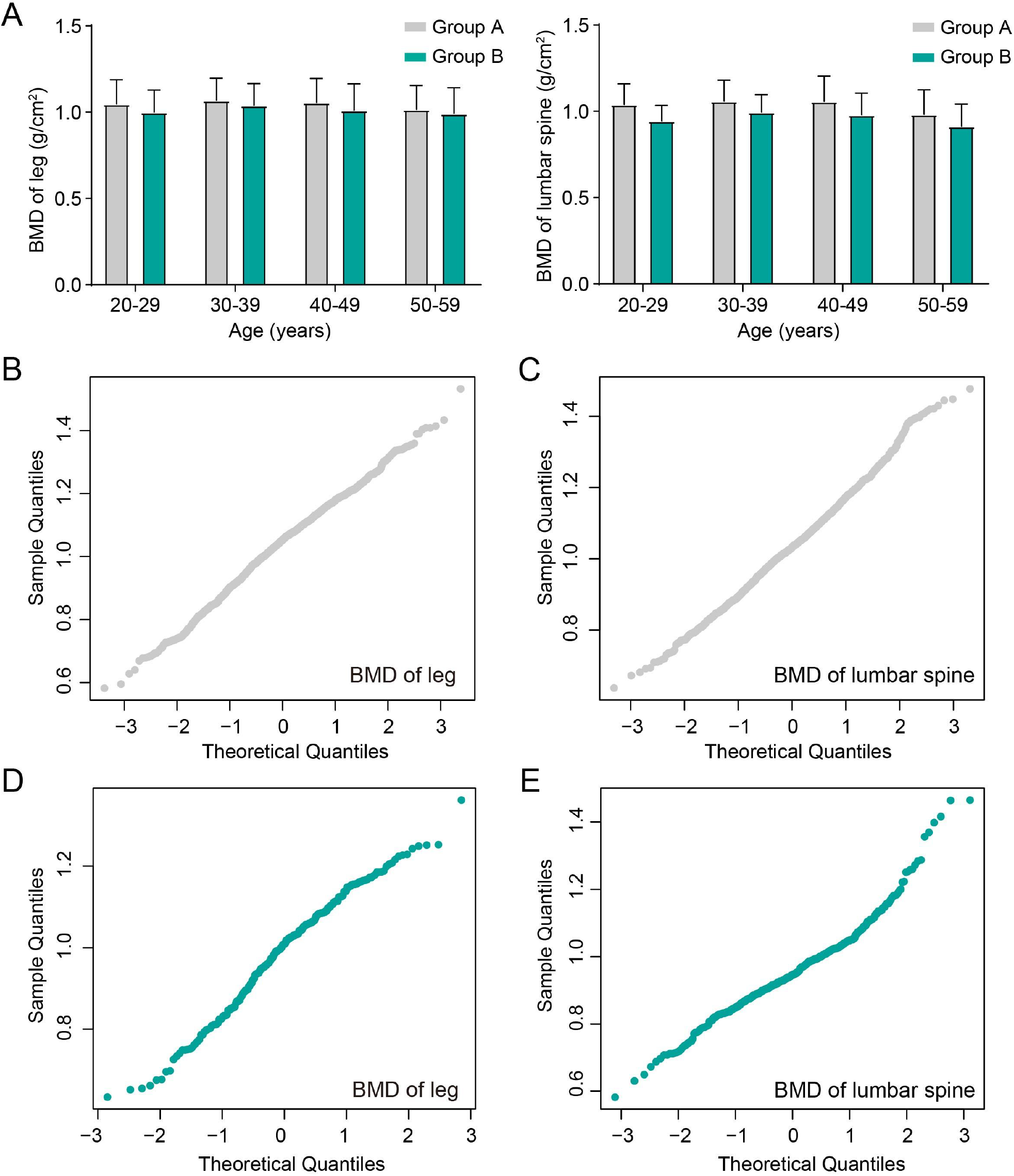
After matching, the bone mineral density (BMD) of leg and lumbar spine in different age groups (A). The normal Q-Q plots of BMD in group A (B, C) and group B (D, E).

### 3.3 Lower BMD in subjects with HPV infection

Welch two-sample t tests showed a lower BMD in subjects infected with HPV in both the legs and lumbar spine (P < 0.001). The decrease in BMD was greater in the lumbar spine than in the legs (Table 1). We performed RCS analysis to more clearly elucidate the association between HPV infection and BMD. In Fig. 2, we found a nonlinear association between the number of cooccurring HPV types and BMD (P for nonlinear association < 0.001). The larger the number of cooccurring HPV types, the lower the BMD was.

**Figure 2.**
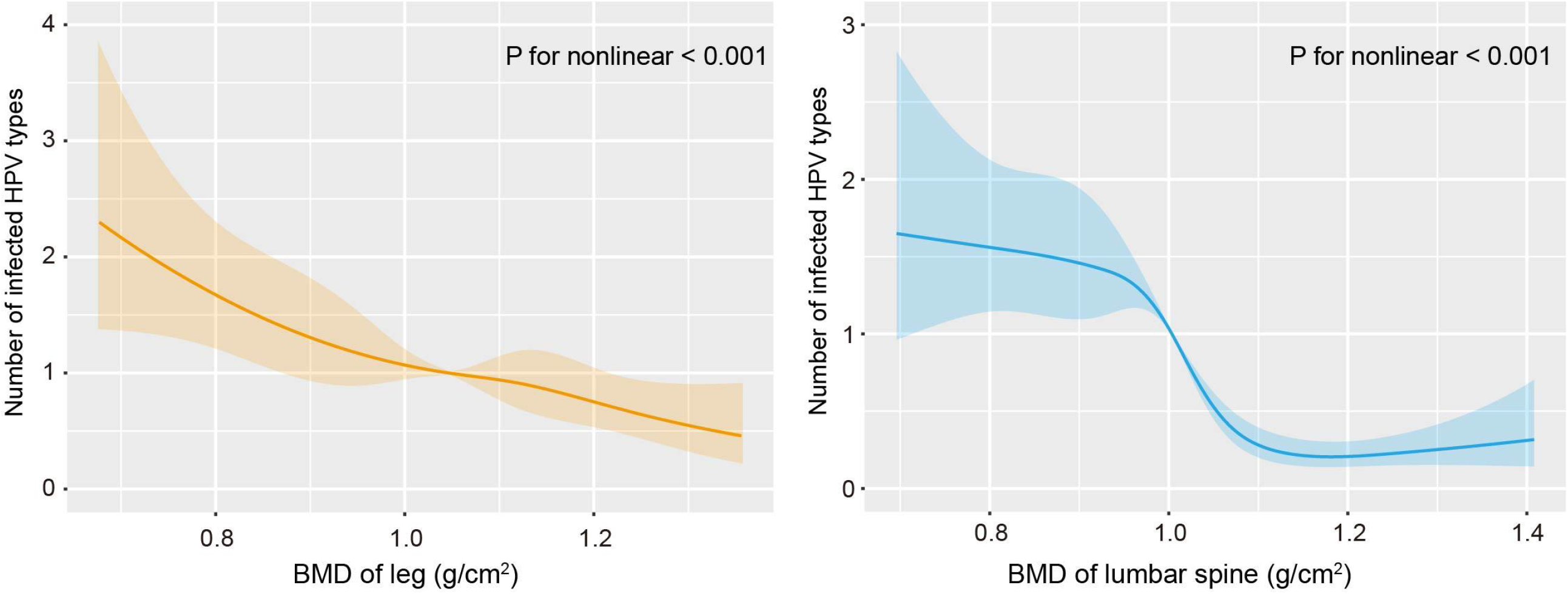
Restricted cubic spline models for the relationship between the number of infected HPV types and bone mineral density (BMD) after matching.

Finally, we analyzed each HPV type in an attempt to identify those with the greatest impact on BMD. The data showed that four HPV types (06, 53, 59 and 89) were negatively correlated with leg BMD (P < 0.05). The strongest correlation was found for type 89 (r = -0.060). Fourteen HPV types were negatively correlated with lumbar spine BMD. Among them, type 53 had the strongest correlation (r = -0.085). There were three HPV types that had an effect on both leg and lumbar spine BMDs, namely, 53, 59 and\ 89. More detailed data are shown in Tables 2 and 3.

## 4. Discussion

To date, the literature on the link between viral infection and OP is scarce. Viruses that have been shown to be associated with OP include human immunodeficiency virus (HIV), hepatitis B virus, hepatitis C virus, and severe acute respiratory syndrome coronavirus 2 (SARS-CoV-2)[36-40]. In the present study, we demonstrated that HPV infection led to a decrease in BMD. Combining the results of the PSM and RCS analyses, we concluded that the association between the two showed a nonlinear negative correlation. The larger the number of cooccurring HPV types, the lower the BMD was.

As mentioned earlier, we confirmed the association between HPV and BMD. However, it is important to determine the causal relationship between them. It often takes a long time for BMD to decline and eventually progress to OP. Compared to this process, HPV infection can be considered an immediate event. Therefore, we believe it is more likely that HPV infection caused the decline in BMD rather than the reverse. In addition, the results after statistical adjustment support our view. PSM eliminated selection bias, prevented our findings from being confounded by other factors, and improved the level of evidence.

However, the biological mechanisms underlying the decline in BMD due to HPV remain unclear. It is known that persistent HPV infection triggers a range of responses, including immunosuppression and abnormal cell destruction[41]. Scott et al. conducted a prospective study and found increased IL-12 concentrations in cervicovaginal lavage fluid specimens in women with persistent HPV infection[42]. This may reflect IL-12-induced macrophage activation and associated subsequent chronic inflammation[43]. The tissue damage caused by inflammation is self-explanatory. In addition, cervical HPV infection also causes infiltration of macrophages and dendritic cells. The activation of these cells also leads to the release of TNF, IL-8 and IL-12[44, 45]. In addition to effects on the immune system, effects of HPV on lipid metabolism have also generated concerns. Lipid metabolism (digestion, uptake, synthesis and peroxidation) is crucial in high-risk HPV infection. The lipid component of the cell membrane makes HPV infection more efficient[46]. Oxidative stress and lipid peroxidation induced by HPV infection also promote cellular damage that may lead to cell death[47]. Although these effects are more pronounced in HPV-infected local tissues, prolonged effects likely lead to systemic damage, including bone damage.

We also noted a larger decline in BMD in the lumbar spine than in the legs in HPV-infected individuals. Fourteen HPV types were negatively associated with lumbar spine BMD, and this number was much higher than that associated with the legs (four types). This finding suggests that lumbar spine BMD is more sensitive than leg BMD. This finding is consistent with the findings of some previous studies to some extent. Snyder et al. found that testosterone increased BMD in older men and that the increase in spinal BMD was larger than that in the hip[48].

Finally, we would like to discuss HPV types 53, 59 and 89. All three types were negatively associated with BMDs in the lumbar spine and legs. Public HPV preventive measures and the detection of HPV infection should additionally focus on these three HPV types. These three HPV types could also help reveal the biological mechanisms underlying the association of HPV with BMD.

Although our statistical analysis attenuated selection bias, some limitations remain in the present study. First, prospective evidence is still needed to truly determine the causal association between HPV infection and BMD decline. Second, elucidating the biological mechanisms of the association between HPV and BMD may require additional molecular experiments. Third, only 37 HPV types were included in this study. However, there are many more types of HPV in existence. How HPV types not detected in the NHANES affect bones health in women is still unknown.

In conclusion, we used data from a nationally representative database of US adults and found a negative association between HPV infection and BMD in female subjects. The larger the number of cooccurring HPV types, the lower the BMD was.

## Contributors

Yunzhen Chen and Guangjun Jiao planned the study and interpreted the analyses. Xiang Li collected data, planned the study, performed the analyses, produced the figures and tables, and drafted the manuscript. All authors contributed to the interpretation of the results and revision of the paper and approved the final manuscript.

## Data Availability

The data generated during and/or analyzed during the current study are available from the corresponding author upon reasonable request.

## Acknowledgments

Not applicable.

---

**Supplemental table 1.** Baseline characteristics for the total subjects before matching.

| Characteristics | Group A<br>N = 2614 | Group B<br>N = 2059 | P value |
| --- | --- | --- | --- |
| <sup>a</sup> Age(years) | 39.00±11.97 | 36.67±12.29 | < 0.001 |
| <sup>a</sup> Poverty income ratio | 2.43±1.76 | 1.91±1.62 | < 0.001 |
| <sup>b</sup> Depression score (<10/≥10) | 1450/269 | 1134/315 | < 0.001 |
| <sup>b</sup> Diabetes (positive/negative) | 126/1933 | 169/2445 | .337 |
| <sup>b</sup> Trouble sleeping (positive/negative) | 172/2439 | 168/1887 | .088 |
| <sup>b</sup> Alcohol consumption (positive/negative) | 712/1811 | 835/1115 | < 0.001 |
| <sup>b</sup> Drinking status (positive/negative) | 130/1732 | 212/1419 | < 0.001 |
| <sup>b</sup> Education level |  |  | < 0.001 |
| Less than 9th grade | 172 | 109 |  |
| 9 - 11th grade | 272 | 285 |  |
| High school graduate/GED or equivalent | 423 | 435 |  |
| Some college or AA degree | 777 | 693 |  |
| College graduate or above | 789 | 378 |  |
| <sup>b</sup> Race |  |  | < 0.001 |
| Mexican American | 456 | 298 |  |
| Other Hispanic | 283 | 219 |  |
| Non-Hispanic White | 1097 | 764 |  |
| Non-Hispanic Black | 379 | 597 |  |
| Non-Hispanic Asian | 58 | 38 |  |
| Other Race | 341 | 143 |  |
| <sup>b</sup> Marital status |  |  | < 0.001 |
| Married | 1457 | 611 |  |
| Widowed | 36 | 49 |  |
| Divorced | 206 | 303 |  |
| Separated | 87 | 103 |  |
| Never married | 459 | 596 |  |
| Living with partner | 190 | 239 |  |

<sup>a</sup> The values were given as the mean and standard deviation.
<sup>b</sup> The values were given as the number of subjects.
P values were calculated by Welch two-sample t test or the Pearson chi-square test with Yates' continuity.

**Supplemental table 2.** Baseline characteristics for subjects of leg BMD after matching.

| Characteristics | Group A<br>N = 1066 | Group B<br>N = 533 | P value |
| --- | --- | --- | --- |
| <sup>a</sup> Age(years) | 39.21±11.25 | 39.93±11.48 | .233 |
| <sup>a</sup> Poverty income ratio | 2.50±1.65 | 2.37±1.67 | .144 |
| <sup>b</sup> Depression score (<10/≥10) | 870/196 | 428/105 | .527 |
| <sup>b</sup> Diabetes (positive/negative) | 73/993 | 39/494 | .729 |
| <sup>b</sup> Trouble sleeping (positive/negative) | 103/963 | 62/471 | .222 |
| <sup>b</sup> Smoking status (positive/negative) | 449/617 | 228/305 | .802 |
| <sup>b</sup> Alcohol consumption (positive/negative) | 449/617 | 228/305 | .802 |
| <sup>b</sup> Education level |  |  | .525 |
| Less than 9<sup>th</sup> grade | 44 | 27 |  |
| 9 - 11<sup>th</sup> grade | 137 | 77 |  |
| High school graduate/GED or equivalent | 198 | 109 |  |
| Some college or AA degree | 369 | 169 |  |
| College graduate or above | 318 | 151 |  |
| <sup>b</sup> Race |  |  | .247 |
| Mexican American | 141 | 82 |  |
| Other Hispanic | 113 | 60 |  |
| Non-Hispanic White | 528 | 272 |  |
| Non-Hispanic Black | 176 | 69 |  |
| Non-Hispanic Asian | 21 | 8 |  |
| Other Race | 87 | 42 |  |
| <sup>b</sup> Marital status |  |  | .345 |
| Married | 520 | 247 |  |
| Widowed | 26 | 13 |  |
| Divorced | 108 | 71 |  |
| Separated | 48 | 21 |  |
| Never married | 252 | 135 |  |
| Living with partner | 112 | 46 |  |

BMD, bone mineral density.
<sup>a</sup> The values were given as the mean and standard deviation.
<sup>b</sup> The values were given as the number of subjects.
P values were calculated by Welch two-sample t test or the Pearson chi-square test with Yates' continuity.

**Supplemental table 3.**
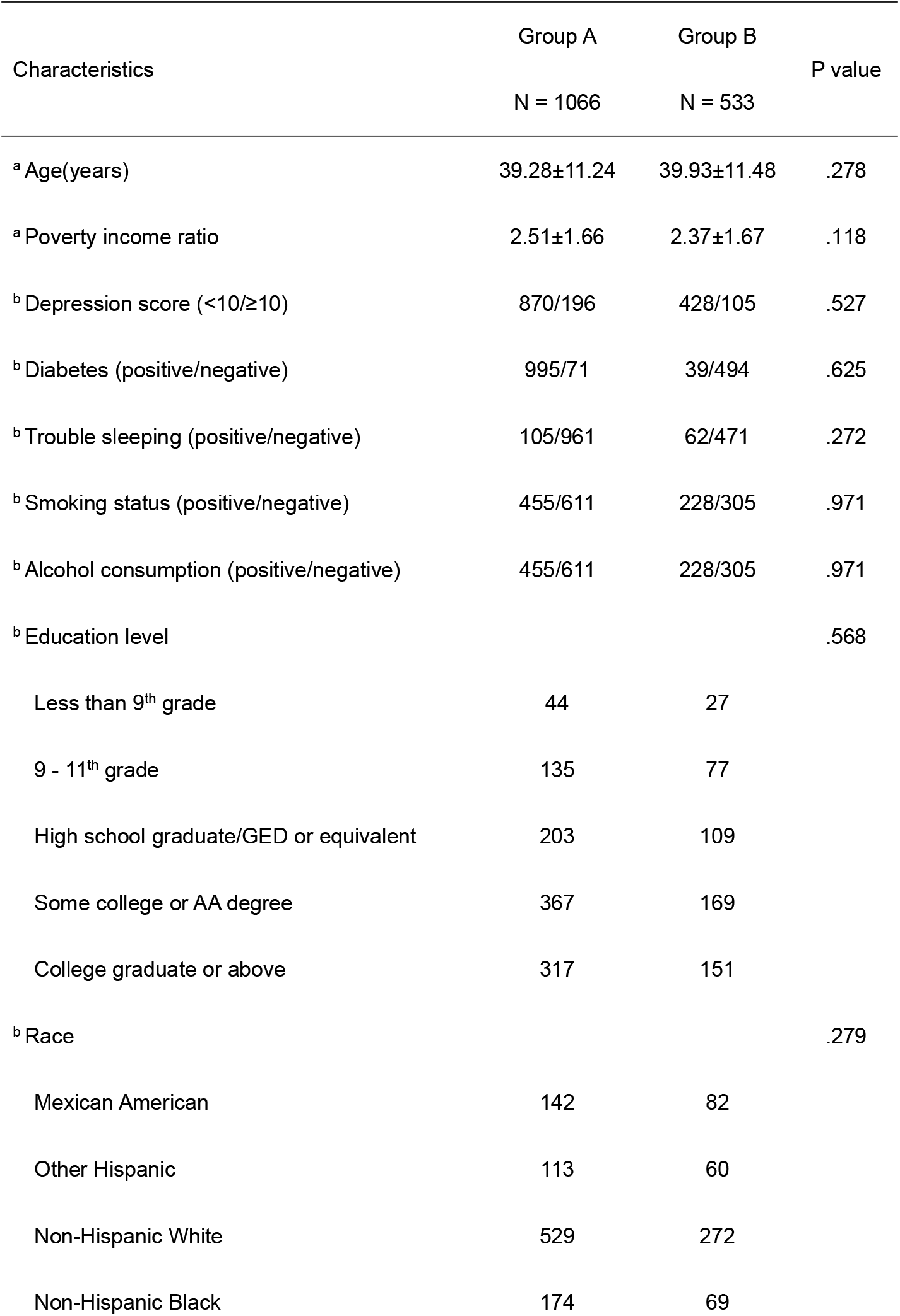
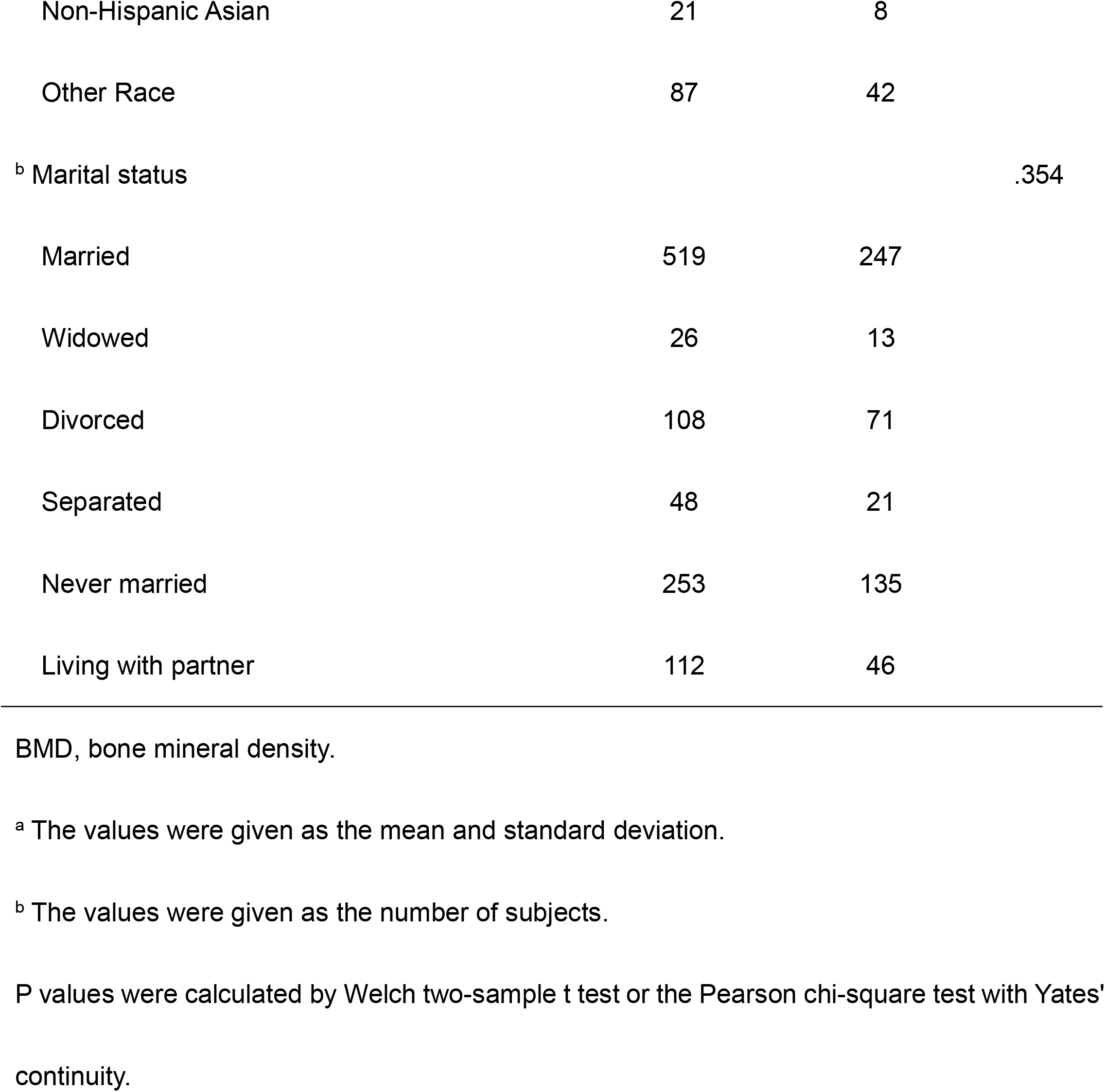
Baseline characteristics for subjects of lumbar.

| Characteristics | Group A<br>N = 1066 | Group B<br>N = 533 | P value |
| --- | --- | --- | --- |
| <sup>a</sup> Age(years) | 39.28±11.24 | 39.93±11.48 | .278 |
| <sup>a</sup> Poverty income ratio | 2.51±1.66 | 2.37±1.67 | .118 |
| <sup>b</sup> Depression score (<10/≥10) | 870/196 | 428/105 | .527 |
| <sup>b</sup> Diabetes (positive/negative) | 995/71 | 39/494 | .625 |
| <sup>b</sup> Trouble sleeping (positive/negative) | 105/961 | 62/471 | .272 |
| <sup>b</sup> Smoking status (positive/negative) | 455/611 | 228/305 | .971 |
| <sup>b</sup> Alcohol consumption (positive/negative) | 455/611 | 228/305 | .971 |
| <sup>b</sup> Education level |  |  | .568 |
| Less than 9<sup>th</sup> grade | 44 | 27 |  |
| 9 - 11<sup>th</sup> grade | 135 | 77 |  |
| High school graduate/GED or equivalent | 203 | 109 |  |
| Some college or AA degree | 367 | 169 |  |
| College graduate or above | 317 | 151 |  |
| <sup>b</sup> Race |  |  | .279 |
| Mexican American | 142 | 82 |  |
| Other Hispanic | 113 | 60 |  |
| Non-Hispanic White | 529 | 272 |  |
| Non-Hispanic Black | 174 | 69 |  |
| Non-Hispanic Asian | 21 | 8 |  |
| Other Race | 87 | 42 |  |
| <sup>b</sup> Marital status |  |  | .354 |
| Married | 519 | 247 |  |
| Widowed | 26 | 13 |  |
| Divorced | 108 | 71 |  |
| Separated | 48 | 21 |  |
| Never married | 253 | 135 |  |
| Living with partner | 112 | 46 |  |

BMD, bone mineral density.
<sup>a</sup> The values were given as the mean and standard deviation.
<sup>b</sup> The values were given as the number of subjects.
P values were calculated by Welch two-sample t test or the Pearson chi-square test with Yates' continuity.

